# Integrated Molecular Profiling of Rhabdomyosarcoma Subtypes by Targeted RNAseq

**DOI:** 10.1101/2024.10.11.24315314

**Authors:** Matthew R Avenarius, Ashley Patton, Nehad Mohamed, Xiaokang Pan, Dan Jones

## Abstract

All subtypes of rhabdomyosarcoma (RMS) show skeletal muscle differentiation but each has different oncogenic mechanisms including recurrent *PAX3* and *PAX7* gene fusions (alveolar), a hotspot mutation in *MYOD1* (spindle/sclerosing) and mutations in other oncogenes and tumor suppressors (embryonal, pleomorphic). This range of genomic findings typically requires several different DNA and RNA-based diagnostic assays. Here, we utilize a clinically validated, targeted RNA next-generation sequencing (RNAseq) panel to simultaneously detect *MYOD1* transcript levels for lineage assignment, *PAX3* and *PAX7* translocations and mutations in *MYOD1* and other RMS-related genes for definitive subtyping in a single assay. RNA-based detection of *MYOD1* p.L122R and other mutations were orthogonally validated with digital droplet (dd)PCR or DNA-based NGS. Expanding the utility of clinical-grade RNAseq data beyond the detection of gene fusions is a cost-effective and time-efficient approach for more comprehensive screening of RMS.

**Impact statement:** Comprehensive and rapid molecular diagnostics is a key driver in the selection and initiation of targeted therapies.

## Introduction

Rhabdomyosarcoma (RMS), a high-grade malignancy derived from skeletal myoblast-like cells, most commonly presents in children (∼7%) and less frequently in adults (∼1%) [1]. The alveolar (ARMS) and embryonal (ERMS) histologic subtypes predominate in children with pleomorphic and spindle cell/sclerosing subtypes more common in adults [2]. The histologic subtypes are correlated with distinct genomic findings including recurrent fusion genes (i.e., *PAX3::FOXO1, PAX7::FOXO1, PAX3::AFX*) in ARMS, aneuploidy and/or recurrent hotspot gene mutations (*TP53, PIK3CA, FGFR4, BCOR* or RAS pathway) in ERMS and pleomorphic subtypes, and a gain-of-function mutation in *MYOD1* NM_002478.4:c.365T>G (p.L122R) and activating *PIK3CA* variants in adult spindle cell RMS and pediatric and adult sclerosing RMS [3,4].

Given the preponderance of fusion genes in soft tissue tumors, anchored multiplex PCR or targeted capture probe-based RNA sequencing (RNAseq) protocols have become the standard molecular diagnostic assays for this tumor type [5,6,7]. However, this will yield a negative result in many soft tissue tumors whenever fusion genes are not the oncogenic driver or when inadequate sampling may have occurred. To address these issues, our clinical laboratory designed a targeted RNAseq fusion panel but supplemented the design with markers of cell differentiation and commonly mutated oncogenes and tumor suppressors. For example, in RMS, the *MYOD1* transcription factor, which is highly expressed in skeletal muscle, was included to assess for rhabdomyomatous differentiation in all tumors tested [8]. To date, this 190-gene RNA-based NGS panel has yielded diagnostic fusions in ∼30% of clinical cases tested at our institution. Here, we investigate the utility of this panel for tumor lineage assessment and mutation detection, using RMS subtyping as a model tumor type.

## Materials & Methods

From the onset of targeted RNAseq panel testing (April 2022) to present, we queried the Molecular Pathology Laboratory database for cases with a final diagnosis of rhabdomyosarcoma (n=10). A similarly sized cohort consisting of 11 high-grade sarcomas without desmin/myogenin expression were also selected as a control for non-muscle derived tumors. The study parameters, including the case series described here, were approved by The Ohio State Cancer Institutional Review Board (Study ID: 2023C0206).

Using a hybrid-capture strategy, a targeted RNAseq panel was designed to capture the full coding regions of 190 genes including RMS-associated mutation targets such as *BCOR, FGFR4, KRAS, MYOD1, NF1, NRAS, PIK3CA, RB1* and *TP53*. The protocol involved initial rRNA depletion and dual-index library construction (Roche, catalog no. KR1151), hybrid capture using custom xGen gene probes (IDT) and sequencing on the Illumina NextSeq 500/550 instrumentation. Base-calling, demultiplexing and conversion of FASTQ sequences were performed using BaseSpace (Illumina). Raw sequence reads were mapped to the human genome (GRCh37/hg19) to generate aligned BAM files using STAR [9]. Fusion detection was performed using Arriba (version 2.40) with annotation and visualization of the data performed by the R script-based program *draw_fusions*.*R* [10]. Variants in genes of interest were identified by manual inspection of the aligned BAM files in IgV (version 2.8.0) [9]. In this study, 400 tumors, including 191 soft tissue tumors were tested with the panel to identify RMS cases.

Oncogene hotspot mutations were confirmed by parallel DNA-based sequencing. For DNA-based NGS panel sequencing, DNA was fragmented by ultrasonicator (Covaris) and libraries were prepared (KAPA HyperPrep). Pre-capture, libraries were amplified and subjected to hybridization of 565 genes (coding sequence) recurrently mutated in cancer. Captured products were again amplified with final libraries sequenced using the NextSeq 500/550 instrumentation.

For *MYOD1* mutation verification, a targeted *MYOD1* NM_002478.4:c.365T>G (p.L122R) DNA-based ddPCR assay was developed. Using 100ng of DNA, the ddPCR reaction consisting of 1.25 μL of 10 μM primers (forward: 5’-CAAGACCACCAACGCCGA-3’; reverse: 5’-CTGGTTTGGATTGCTCGACG-3’), 0.625 μL of 10 μM probes (wild type: HEX-CCGC+C+T+GAGCAAA; mutant: FAM- CGC+C+G+GAGCAAA), and 11 μL 2x ddPCR Supermix for probes-No dUTP (Biorad) was assembled. The BioRad QX200 instrumentation was used for droplet generation and analysis. The companion QX Manager software (version 1.2) was used to calculate *MYOD1* NM_002478.4:c.365T>G (p.L122R) mutation levels. The assay was validated with additional RMS-cases at a sensitivity level of approximately 0.2% mutated allele based on dilution series (data not shown).

## Results

Among the 400-tumor set, elevated *MYOD1* transcripts levels were only seen in cases diagnosed as RMS (alveolar=1, sclerosing/spindle cell=4, cutaneous epithelioid=1, pleomorphic=3) based on morphology and immunohistochemical positivity for desmin and/or myogenin, and in one high-grade neoplasm with focal rhabdomyosarcomatous differentiation. In Figure 1A, mean NGS reads across all coding exons of *MYOD1* were compared for these 10 RMS cases to a control group of 11 high-grade sarcomas without desmin/myogenin expression. Above 800 *MYOD1* reads per sample, there was 100% specificity between *MYOD1* transcript levels and immunohistochemical evidence of skeletal muscle differentiation. One RMS case showed <800 *MYOD1* reads, which was likely due to focal expression of skeletal muscle markers (Figure 1E).

**Figure 1.**
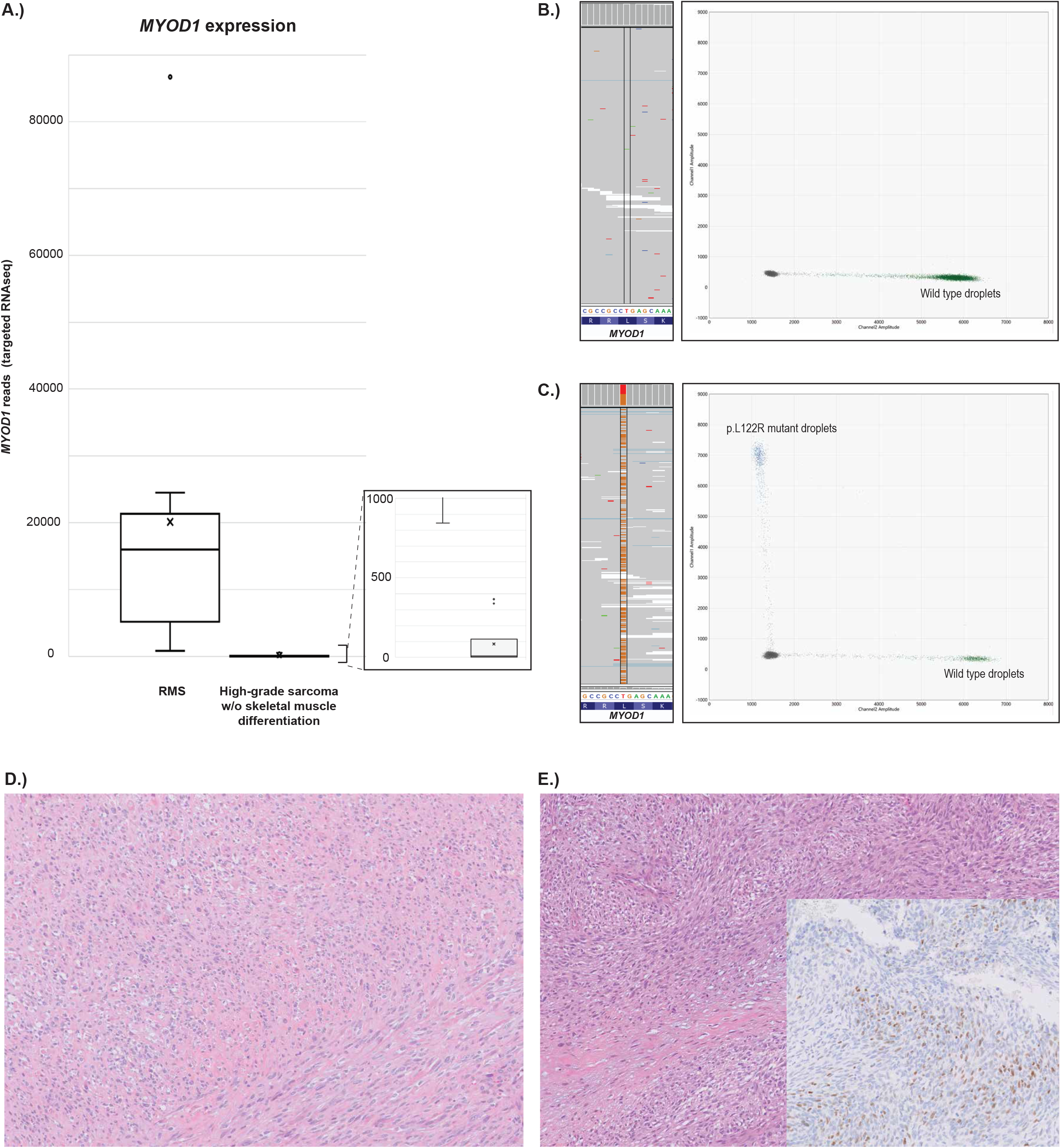
*MYOD1* expression and mutation calling (p.L122R) from targeted RNAseq data. A.) *MYOD1* expression levels as determined by mean read depth from targeted RNAseq data in RMS and controls (other high-grade sarcomas). B.) IGV visualization of targeted RNAseq data and orthogonal ddPCR data for rhabdomyosarcoma *MYOD1* p.L122R (c.365T>G) negative specimen. C.) IGV visualization of targeted RNAseq data (*MYOD1* c.365 p.L122R read depth = 5456; VAF=54) and orthogonal ddPCR data (VAF=42.9) for RMS *MYOD1* p.L122R (c.365T>G) positive specimen (case 1). D.) *MYOD1* c.365 p.L122R-mutated sclerosing/spindle cell RMS (case 11, 200x magnification). E.) Malignant peripheral nerve sheath tumor with focal MYOD1, desmin and myogenin expression (insert) and mid-level *MYOD1* transcripts (case 12).

One case of alveolar RMS with *PAX3::FOXO1* was identified by the fusion portion of the comprehensive RNAseq assay. To subtype fusion-negative RMS, *MYOD1* reads were examined for evidence of the RMS- associated NM_002478.4:c.365T>G (p.L122R) mutation. Two spindle cell/sclerosing RMS cases showed the *MYOD1* p.L122R mutation at variant allele fractions of 54% and 47% (case 1, 11). *MYOD1* was not included on the concurrent DNA-based NGS study; therefore, a DNA-based ddPCR assay for the *MYOD1* p.L122R mutation was developed. This assay confirmed the presence of the p.L122R mutation in cases 1 (VAF=49.5%) and 11 (VAF=42.9%) and was negative in all other cases included in this study (Figure 1B-C).

Accuracy of mutations detected across the full 190-gene set in the RNAseq assay was assessed by comparison with DNA-based NGS, particularly in the fusion-negative/*MYOD1* mutation-negative cases. The RNAseq assay detected hotpot, frameshift and nonsense mutations in the *NF1, PTEN, RB1* and *TP53* tumor suppressor genes at a similar fraction, in a total of 7 calls across 4 tumors (Table 1). One inactivating *NUTM1* mutation seen by DNA-NGS was not detected by RNAseq due to loss of that transcript in the tumor (case 10).

**Table 1.**
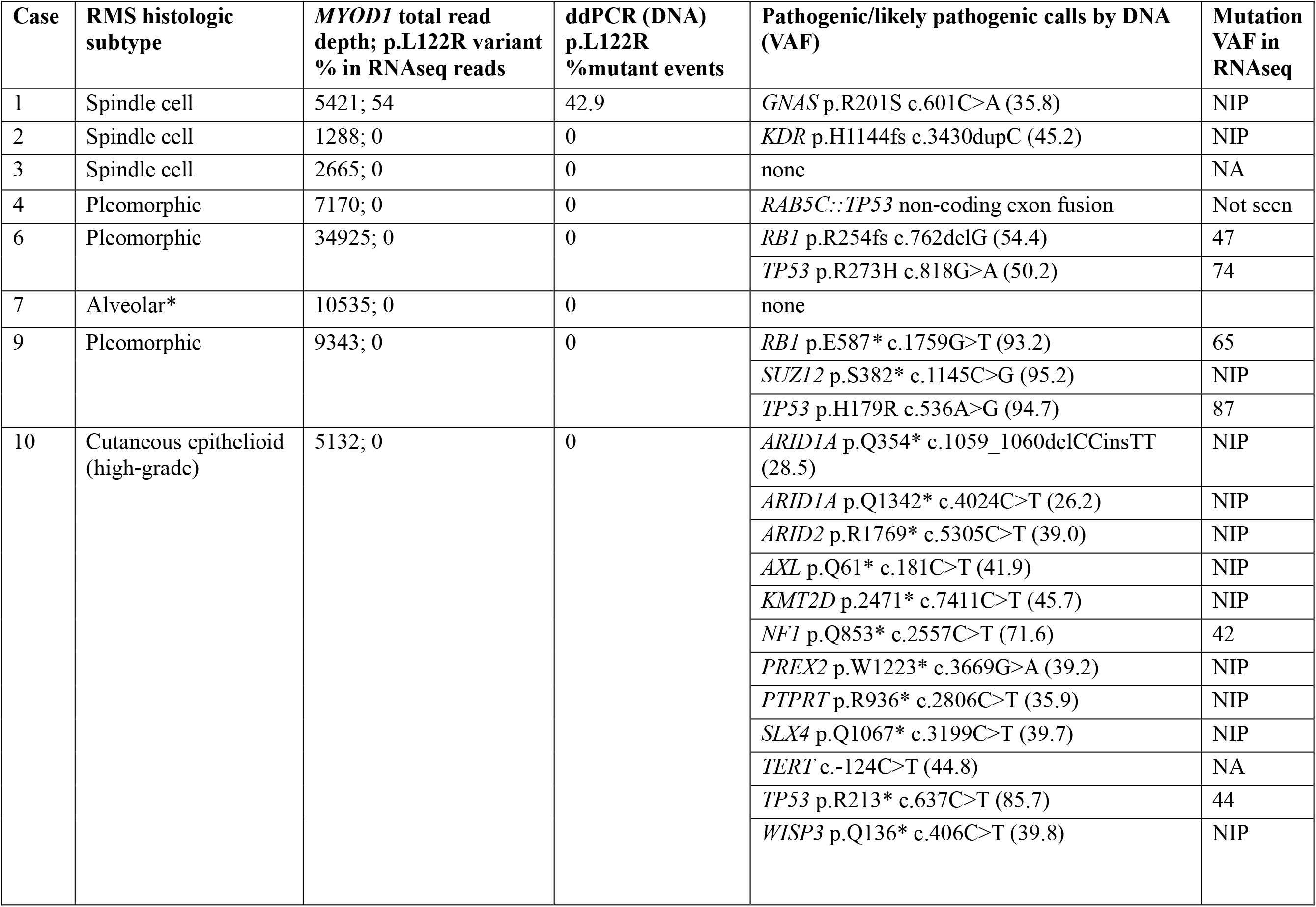
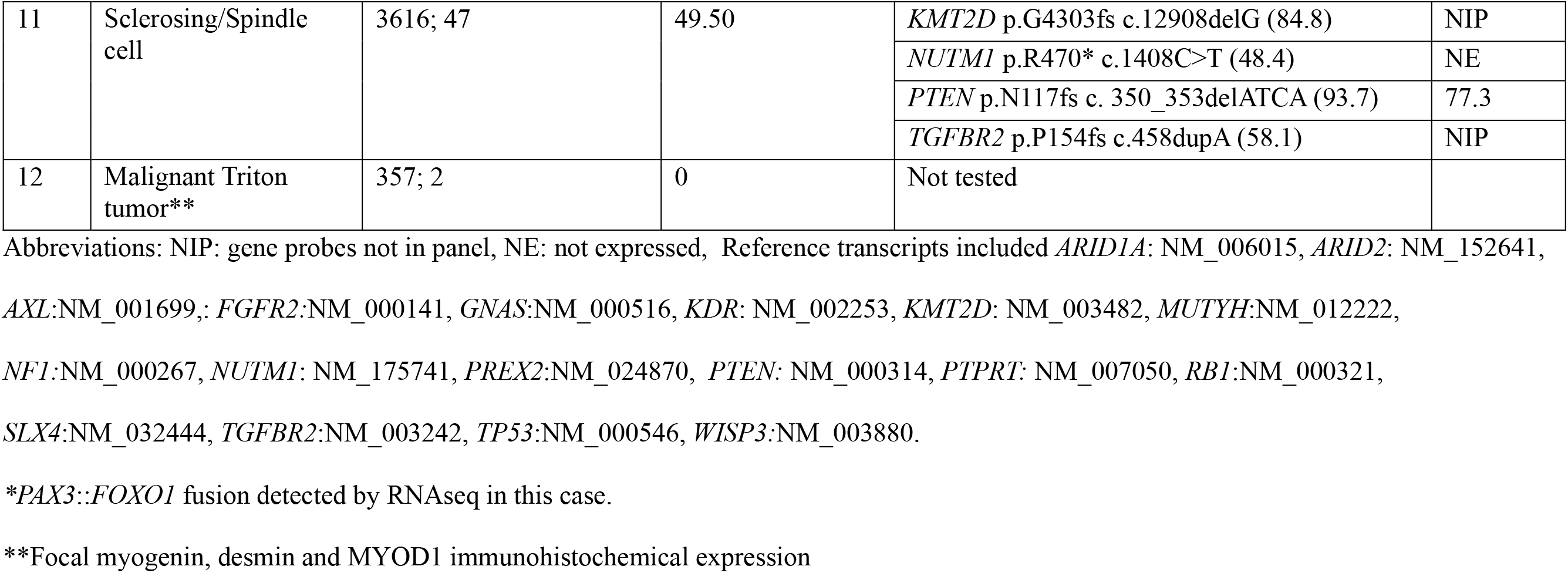
Comparing RNAseq, DNA NGS and *MYOD1* ddPCR for molecular profiling of rhabdomyosarcomas.

## Discussion

We show the design of a targeted RNAseq assay to include transcript markers of tumor lineage and common mutations, particularly *MYOD1* NM_002478.4:c.365T>G (p.L122R) and tumor suppressor loci, can capture the genetic heterogeneity of RMS subsets and introduces efficiency into the molecular diagnostic workup. The benefit of including lineage-markers in RNAseq diagnostic panels is validated here for *MYOD1*. Partial skeletal muscle differentiation is mostly seen in pleomorphic RMS/carcinosarcoma, with one case included here showing correlation of that finding with only mildly elevated *MYOD1* transcript levels. Although detecting skeletal differentiation by immunostaining is straightforward given the known high specificity of markers like myogenin, this diagnosis can be missed/overlooked in more poorly differentiated tumors.

Mutation detection by RNAseq represents another aspect of the clinical panel utilized here. In two of the spindled RMS cases studied, the *MYOD1* NM_002478.4:c.365T>G (p.L122R) mutations detected by RNAseq were concordant between targeted RNAseq data and the orthogonal ddPCR assay. Integrating mutation calling into our standard gene fusion workflow allows for rapid and accurate assessment of RMS including *MYOD1* expression levels to verify the origin of the tumor, fusion gene analysis, and *MYOD1* p.L122R mutation calling. This strategy eliminates the need to order additional stand-alone *MYOD1* c.365T>G (p.L122R) testing that may contribute to delays in case sign out pending completion of comprehensive genomic panels. Despite the potential for nonsense mediated decay selective loss of transcripts due to inactivating variants (as seen for one *NUTMI1* call), the five nonsense/frameshift mutations seen in common tumor suppressor genes were well-retained here in the RNAseq data. While the scope of this study is focused on broadening the utility of our targeted RNAseq data for the diagnosis of RMS, this strategy may be adapted for other tumor types with emphasis on the importance of orthogonal validation of mutation calling.

## Data Availability

All data produced in the present work are contained in the manuscript

## Acknowledgements

We acknowledge Nikkie Nguyen, Harish Muniyappa, and Katherine Dunkel for preparing libraries and sequencing the specimens included in this manuscript. The laboratory of Christopher Corless, Oregon Health Sciences University, provided additional RMS cases for the ddPCR assay validation. The authors received no specific funding for this work.

## Statement of author contributions

M.R.A. and D.J. performed study concept and design; A.P., S.E.L, N.M., X.P., performed development of methodology; M.R.A. and D.J. prepared and edited manuscript. All authors read and approved the final paper.

## Ethical approval

This study was reviewed and approved by The Ohio State Cancer Institutional Review Board (Study ID: 2023C0206).

